# Has the design quality of randomized controlled trials of Acupotomy improved over the past 18 years? -- CONSORT statement-based literature study from 2006 to 2024

**DOI:** 10.1101/2024.10.15.24315538

**Authors:** Junjie Li, Yantong Zhou, Xinzhu Lu, Ying Bian

## Abstract

Acupotomy, originally named ‘Nine Needles’ in *Ling Shu*, was invented as a new type of TCM instrument in 1976, an innovative combination of acupuncture and surgical treatment. Its forward development is related to the Clinical effectiveness and safety. RCTs are gold standards in clinical practice and were welcomed in Acupotomy recently. The CONSORT Statement is set to guide the designing, analysis and interpretation of trials. But there haven’t been many Acupotomy RCTs until now, with few on quality evaluation, therefore design quality of Acupotomy RCTs is still weakness. This study aims to assess design quality of acupotomy RCTs by CONSORT statement, to analyze the overall quality status and influencing factors.

PubMed database was used to search keywords like ‘Acupotomy’ and ‘Randomized Controlled Trial’. All 48 Acupotomy RCTs published from January 2006 to January 2024 were included. The CONSORT(2010) was used for quality assessment.

48 studies were included for analysis, with 39 articles from Grade 3A hospitals and 9 from non-Grade 3A hospitals. Scores of RCTs ranged from 33 to 82, the mean score of 53.1 and median of 49. Grade 3A and non-Grade 3A hospitals differed significantly only in item 8, no studies reported item 18, and items 11, 14, and 23 had the highest frequency of reporting as failed.

Based on 48 Acupotomy RCTs included, the publication time associated with the quality of reports. The number of authors and possession of funding were the most important factors affecting the total score. Number of beds, hospitals’ grade, sample sizes, and region GDP/PP did not relate to the total score. Among 25 items, Ancillary analyses, Blinding, Recruitment were the worst-performing items. Therefore, updating and standardizing the use of CONSORT can help to improve quality of RCTs, and cross-team communication and cooperation could promote the use of CONSORT.

## Introduction

### Acupotomy

A new-style bladed needle that has a flat head and a cylindrical body is named Acupotomy, which is a particular type of acupuncture, using a blade-needle combined with a flat surgical scalpel at the tip of the needle(1, 2). Through stripping adhesions and releasing contractures of deep soft tissues through the flat knife at the tip of the needle(3), it has been widely used for the treatment of musculoskeletal pain, soft tissue injuries, and bone hyperplasia(4), such as knee osteoarthritis(3, 5).

The Acupotomy was originally derived from ‘The Nine Needles’ recorded in the *Lingshu*(6), then was improved and reinvented by Prof. Zhu Hanzhang in China in 1976(7, 8), and it also been described with similar terminologies such as Acupotomology, Acupotome, needle knife, needle scalpel, miniscalpel, stiletto needle, sword-like needle, mini needle knife and xiaozhendao(9). Acupuncture is one of the most widely recognized Complementary and Alternative Medicine(CAM) therapies(10), and clinical studies have shown that Acupotomy is more effective in improving pressure-pain thresholds(11), and also has the effect of reducing inflammatory factors(12).

As an innovative and organic combination of traditional acupuncture therapy in Traditional Chinese Medicine(TCM) and surgical therapy in Western medicine today(13), Acupotomy shows promise as a new alternative to non-pharmacologic interventions(14), while the clinical effectiveness and safety are the key factors affecting its continued development(15, 16).

### Randomized control trials

Randomized control trials (RCTs) are the gold standards in evaluating and efficiently translating research data into clinical practice(17). Owing to requiring large amounts of patient data to identify modest differences between treatments, a well-designed and rigorously conducted RCT can produce the most valid and precise scientific evidence to a certain extent(18). Thus, considering it could adequately assess the efficacy of an intervention (ie, what can work)(19), RCTs have been welcomed in Acupotomy clinical trials recently, especially represented by the analysis of the specific pain or disease, such as Lumbar Spinal Stenosis(20) and Cervical Spondylosis(21).

However, completing all included trials and results analysis does not equate to finishing a RCT trial at a high level, the quality of its completion is one of the determinative factors in evaluating the trails’ value(22). For instance, Lin has pointed out that partial trials were of low quality and adverse effects were not recorded in s systematic review of the effects of small needle-knife therapy (23).

### The CONSORT statement

To help increase trial utility, replicability and transparency(24), a team of professional editors and scientists developed the CONSORT (Consolidated Standards for Reporting of Trials) statement(17), which has been supported by more than 400 journals and multiple editorial groups(e.g., International Committee of Medical Journal Editors) since its publication in 1996(25, 26), and was widely recognized internationally(27, 28).

However, CONSORT has been adopted relatively late in China (29, 30). Only a few Chinese medical journals recommended using CONSORT in the part of Author Instructions or Guidelines previously(31). In recent years, it has gradually received more attention(32-34).

The CONSORT statement is set to regulate the standards for the trial’s design, analysis and interpretation of the results(35). It’s made up of a 25-item checklist that provides authors with a solid backbone around which to construct and present an RCT(36).

While most Acupotomy clinical researches focus on the systematic review and meta-analysis of RCTs, taking the risk of bias as the indicator for evaluating quality on the whole, there is little study that concentrates on the perspective of CONSORT assessment(37). So, this study aims to assess the design quality of acupotomy RCTs from 2006 to 2024 by using the CONSORT 2010 statement as the evaluation criterion, revealing whether there have been changes in the design quality of acupotomy RCTs over the past 18 years.

## Materials and Methods

### Literature selection

Using the PubMed database, all randomized controlled trials published by January 2024 assessing the efficacy of Acupotomy were included. The search strategy is shown in Table 1.

**Table 1.**
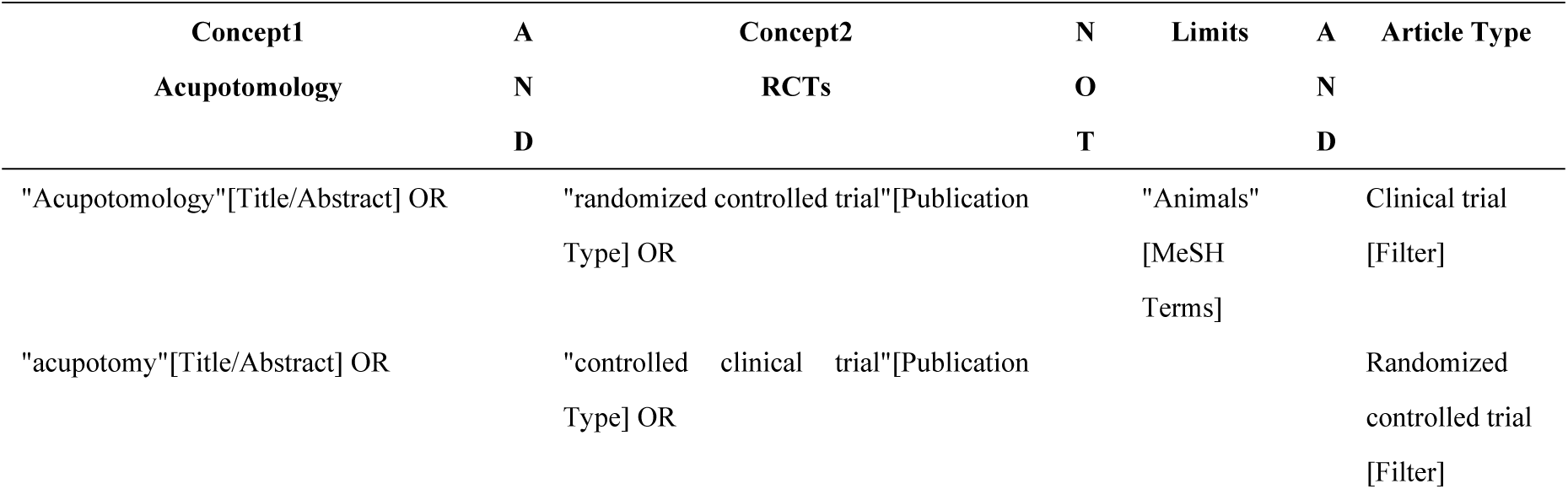
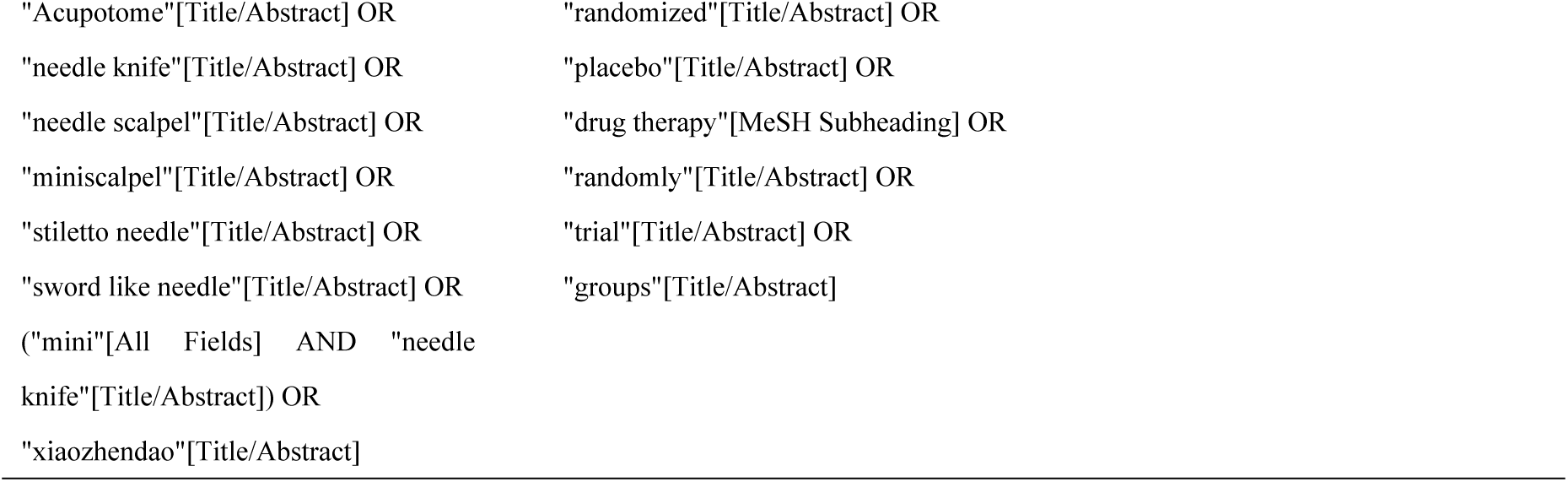
The search strategy.

To confirm study eligibility, searches were followed by a manual review in which study titles were examined first, then abstracts, and finally the full text.

#### Inclusion criteria

Reports were included only if they involved human subjects. There were no language restrictions on the reports. Acupotomy can be used alone or in combination with drugs or other therapeutic measures. There is no restriction on the technique or type of acupotomy, including traditional acupotomy, point acupotomy and other new types of acupotomy.

#### Exclusion criteria

Studies related to Needle-knife Fistulotomy(NK-F), draft study protocols, case-control studies, studies that have not been finalized, systematic reviews, meta-analyses, and follow-up studies of previously published trials.

### Report evaluation criteria

All evaluators assessed articles using the revised CONSORT2010 checklist (Table 2), the CONSORT Interpretation Guidance document, and the examples cited within it. Since the CONSORT list does not assign weights to the 25 items(38, 39), in this study, 25 items were assigned an average of 4 points for a total of 100 points based on the experience of the relevant research literature.

**Table 2.**
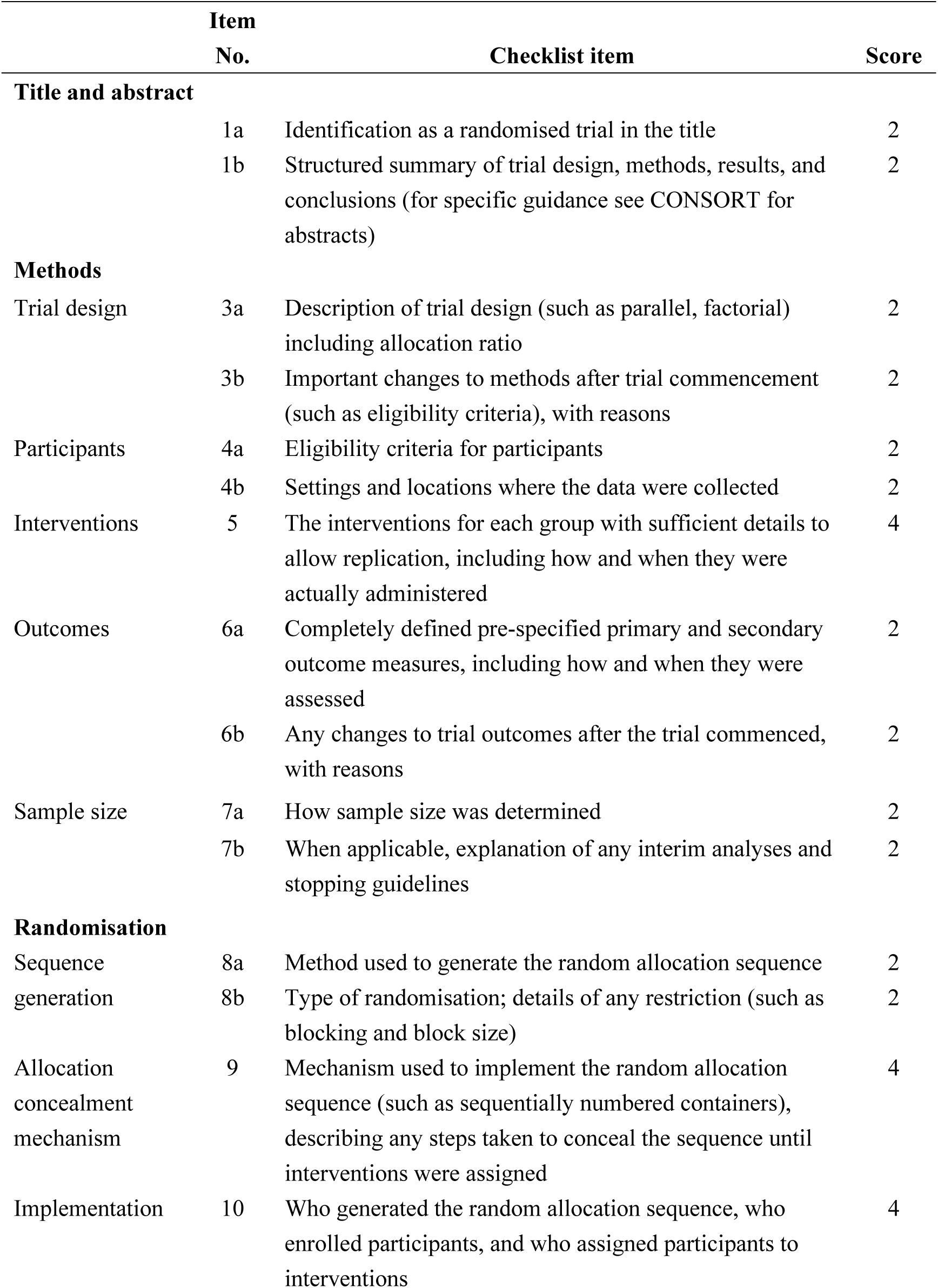
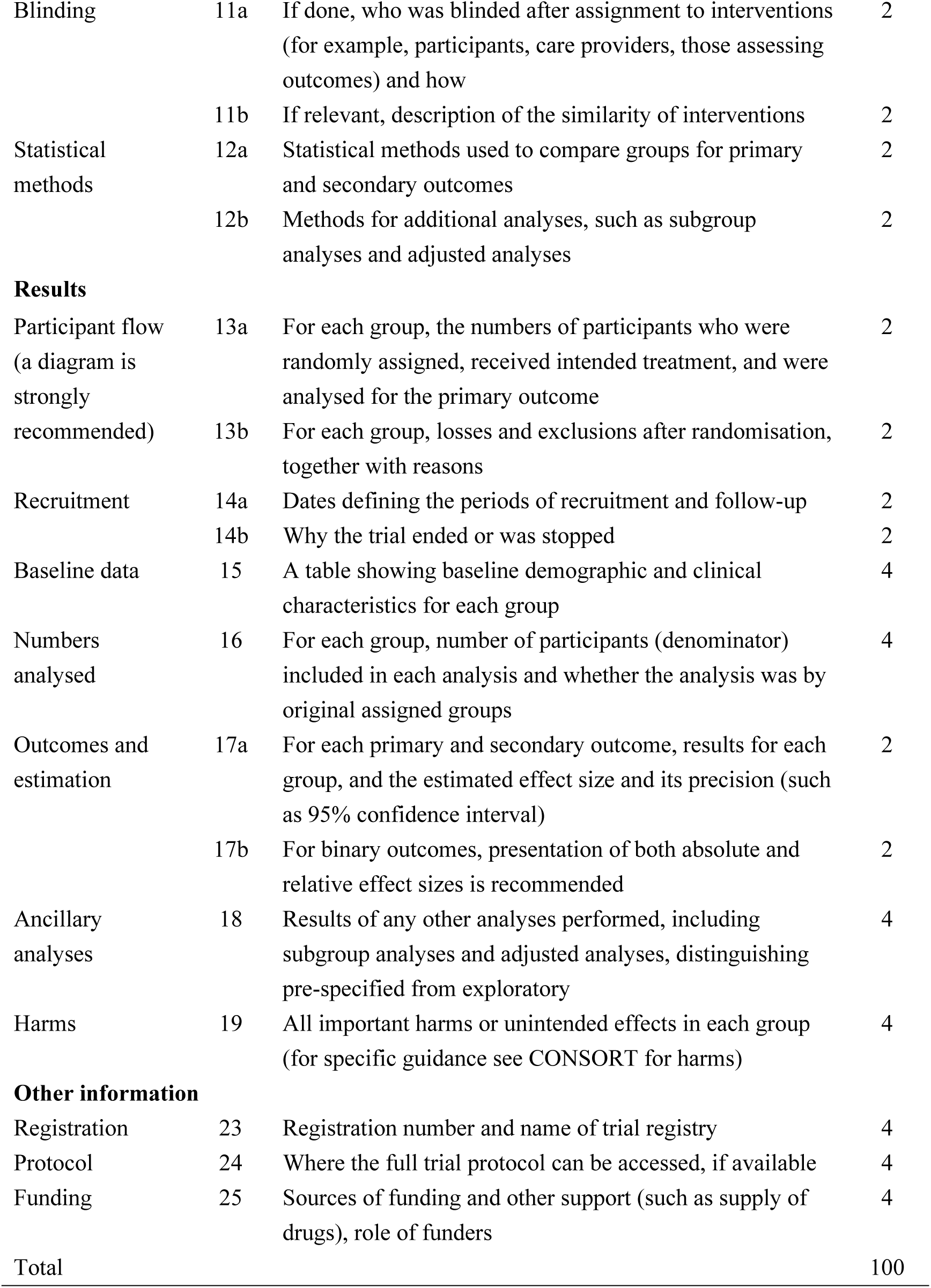
CONSORT 2010 Checklist.

The 2010 revised CONSORT statement has 25 items, 12 of which are divided into two sub-items (37 items in total). 13 individual items without sub-items are awarded 4 points for adequate reporting, 2 points for inadequate reporting or mentioning only, and 0 points for missing items, while 12 items with sub-items are awarded 2 points for each sub-item of adequate reporting, 1 point for inadequate reporting or mentioning only, and 0 points for missing items, and the sum of the two sub-items is the score for this item(40).

A report is considered adequate only if it is detailed explicit and written in the required structural subdivisions. If the content is sufficient but appears in a structure not the CONSORT required, the report is considered insufficient or only mentioned. Additionally, the location of the setting where the data were collected in item4b was judged to be adequate only if the content included, but was not limited to, the hospital, department, time of collection, and time of treatment. Item 15 required a table of baseline data for subjects in the Result, but if the study included a table of baseline data for the population in the section of Method and did not state it in the section of Result, the report was deemed inadequate.

### Data extraction and evaluator training

48 studies were evaluated independently by three evaluators. In addition to the evaluated scores, the extracted data also included descriptive information such as year of publication, sample size, number of authors, level of hospital to which the study belonged and number of established beds, foundation support for the study, and GDP per capita value for 2023 at the municipal level in the region of publication (the region of publication was determined based on the address of the first author’s institution).

The three assessors, all professionals from the University of Macau with a background in medical administration, were trained in assessing RCTs using the CONSORT checklist, and the definition of each checklist item was discussed and agreed upon before the data extraction. The full text of each RCT was then assessed independently by each evaluator. If the inter-assessor concordance was low, a fourth professional assessor was introduced to further assess the full text, and the final agreed score would be confirmed after the consultation in all four.

### Statistical analysis

All data were typed into the extraction form and descriptive statistical data analysis was performed using Microsoft Excel 2007 and SPSS 29. The level of significance was set at P<0.05.

Cohen’s kappa was determined to assess inter-assessor agreement for each CONSORT item. Kappa values between 0.6 and 0.8 were considered to indicate substantial agreement, while values above 0.8 were considered to indicate almost perfect agreement.

The univariate analyses as well as multifactorial impact analyses of possible determinants were performed. The date of study publication and the 2023 GDP per capita (CNY) of the hospital’s city were used as independent variables and the total score as the dependent variable for linear regression analysis. The chi-square test was utilized to study the impact of Grade 3A hospitals versus non-Grade 3A hospitals on the extent of each item. The correlation analysis was used to study the relationship between the number of beds, the number of authors, the sample size and the 25 items. The Pearson analysis was performed to analyze the factors affecting the scores.

## Results

An initial literature search identified 93 studies from October 1994 through January 2024. After the screening of titles and abstracts, 32 studies were excluded as they were irrelevant, including 9 articles from between 1994-2005, all of which were excluded for related cutting techniques such as needle-knife fistulotomy(NK-F). The remaining 61 articles from the beginning of 2006 were evaluated for full text, of which 6 articles were not available to read and 7 were considered not eligible, resulting in the inclusion of 48 studies for analysis. Of the 48 eligible articles, 6 were published before 2010, 42 were published later, 46 published were from China, and 2 were from South Korea; the literature screening process and results are shown in Fig 1.

**Figure 1.**
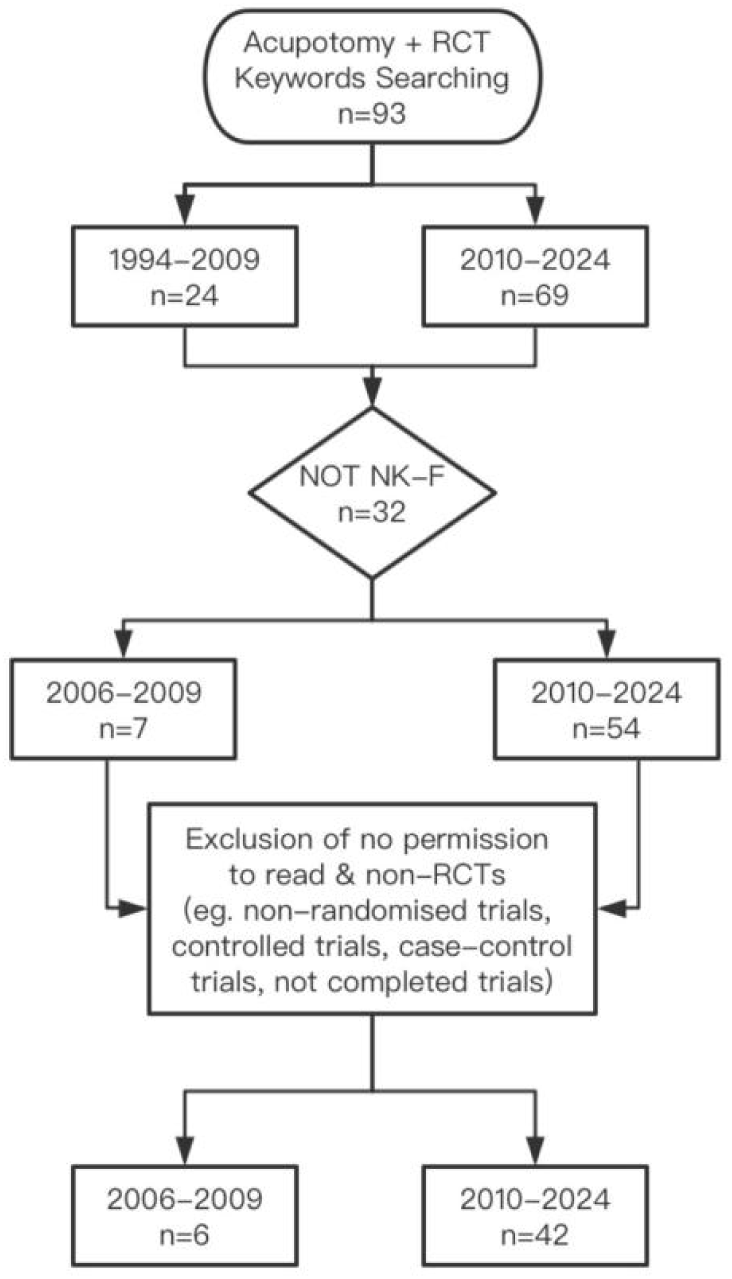
Literature selection flowchart.

### Overall compliance with the CONSORT statement

As shown in Fig 2, the lowest score out of 48 studies from 2006 to 2024 was 33, the highest score was 82, the mean score was 53.1, the median score was 49, and the quality of reporting improved over time. Using the year of publication as the independent variable (X) and the total score as the dependent variable (Y), it was found that there is a correlation between the year of publication and the total score of the report (R^2^=0.455, F=38.350, P<0.001).

**Figure 2.**
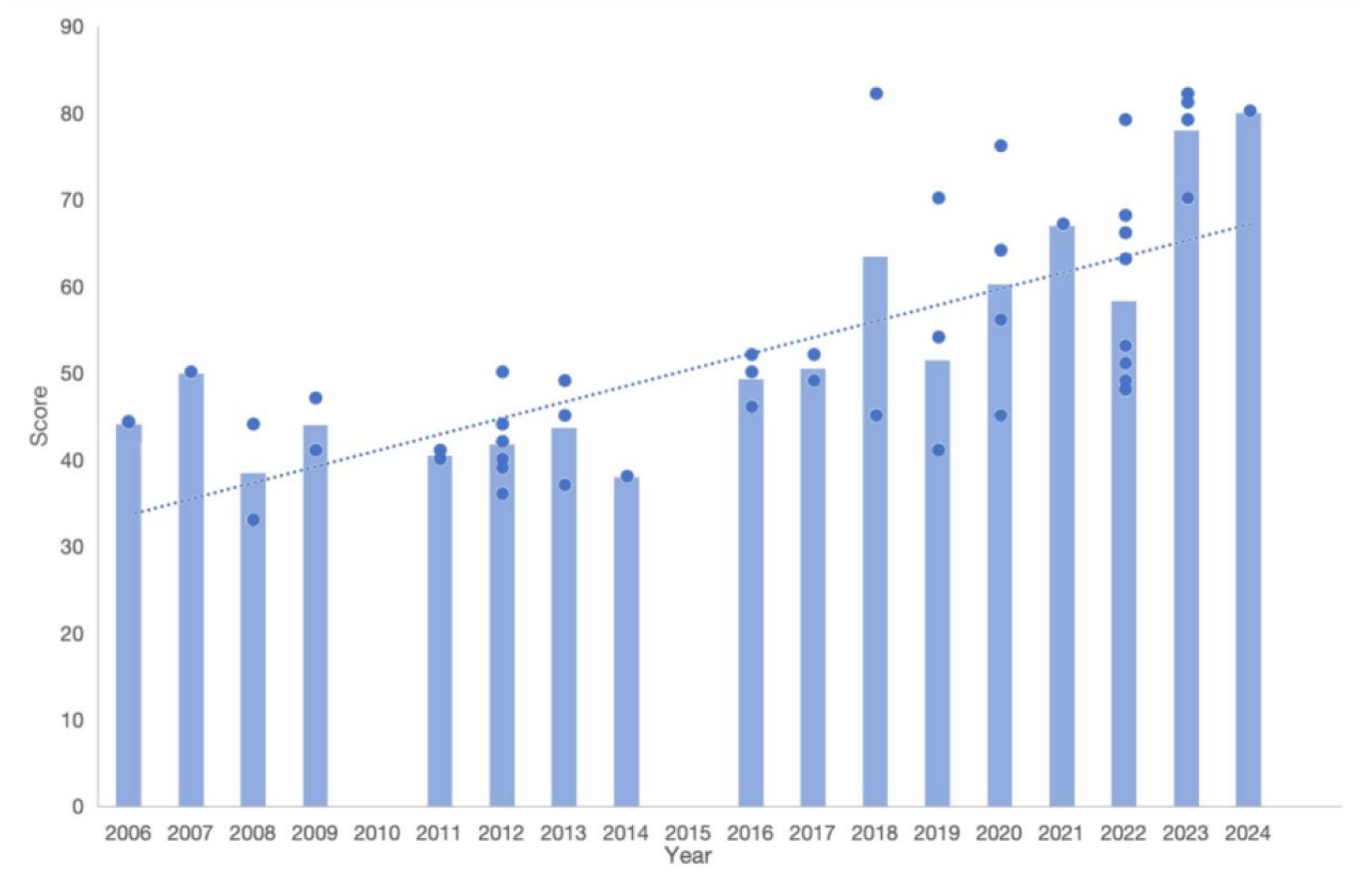
The bar chart with scatter of publication year versus score of report quality.

### Compliance with the CONSORT statement in different grades of hospitals

Of the 48 articles, 39 were sourced from Grade 3A hospitals and 9 from non-Grade 3A hospitals. To compare whether the two hospitals responded differently to items, the mean scores of the RCTs of Grade 3A hospitals and non-Grade 3A hospitals for each of the 25 items were calculated separately in Table 3, as well as the frequency of obtaining excellent, good, and failed, where a score of 0 is considered as failed, 0-2 is considered as accepted, and 3-4 is considered as excellent.

**Table 3.** Mean scores of 25items and their frequency of excellent, accepted and failed.

| Items | Mean<br>(Grade<br>3A) | Mean<br>(Non-Grade 3A) | Grade 3A |  |  | Non-Grade 3A |  |  | X<sup>2</sup> | P |
| --- | --- | --- | --- | --- | --- | --- | --- | --- | --- | --- |
|  |  |  | excell<br>ent | accep<br>ted | failed | excell<br>ent | accep<br>ted | failed |  |  |
| 1 | 2.90 | 2.67 | 18 | 21 | 0 | 3 | 6 | 0 | 0.488 | 0.485 |
| 2 | 3.74 | 3.67 | 39 | 0 | 0 | 9 | 0 | 0 | - | - |
| 3 | 1.97 | 2.00 | 0 | 39 | 0 | 0 | 9 | 0 | - | - |
| 4 | 3.69 | 3.78 | 37 | 2 | 0 | 9 | 0 | 0 | 0.482 | 0.488 |
| 5 | 3.90 | 3.78 | 37 | 2 | 0 | 8 | 1 | 0 | 0.447 | 0.504 |
| 6 | 1.90 | 1.89 | 1 | 38 | 0 | 0 | 9 | 0 | 0.236 | 0.627 |
| 7 | 1.13 | 0.78 | 5 | 13 | 21 | 0 | 4 | 5 | 1.413 | 0.493 |
| 8 | 3.49 | 4.00 | 34 | 3 | 2 | 0 | 8 | 1 | 29.302 | 0.001* |
| 9 | 1.38 | 0.89 | 11 | 5 | 23 | 2 | 0 | 7 | 1.664 | 0.435 |
| 10 | 1.13 | 0.67 | 6 | 11 | 22 | 1 | 1 | 7 | 1.499 | 0.473 |
| 11 | 0.38 | 0.44 | 0 | 10 | 29 | 0 | 3 | 6 | 0.219 | 0.640 |
| 12 | 2.05 | 2.00 | 1 | 38 | 0 | 0 | 9 | 0 | 0.236 | 0.627 |
| 13 | 1.97 | 1.56 | 14 | 11 | 14 | 3 | 2 | 4 | 0.253 | 0.881 |
| 14 | 0.46 | 0.44 | 3 | 5 | 31 | 0 | 3 | 6 | 2.694 | 0.260 |
| 15 | 2.62 | 2.22 | 18 | 14 | 7 | 3 | 5 | 1 | 1.194 | 0.551 |
| 16 | 4.00 | 4.00 | 39 | 0 | 0 | 9 | 0 | 0 | - | - |
| 17 | 2.00 | 2.00 | 0 | 39 | 0 | 0 | 9 | 0 | - | - |
| 18 | 0.00 | 0.00 | 0 | 0 | 39 | 0 | 0 | 9 | - | - |
| 19 | 1.18 | 0.67 | 8 | 5 | 26 | 2 | 0 | 7 | 1.295 | 0.523 |
| 20 | 1.74 | 1.33 | 18 | 0 | 21 | 2 | 0 | 7 | 1.723 | 0.189 |
| 21 | 3.95 | 4.00 | 38 | 1 | 0 | 9 | 0 | 0 | 0.236 | 0.627 |
| 22 | 3.95 | 4.00 | 38 | 1 | 0 | 9 | 0 | 0 | 0.236 | 0.627 |
| 23 | 0.82 | 0.44 | 8 | 0 | 31 | 1 | 0 | 8 | 0.424 | 0.515 |
| 24 | 1.28 | 1.33 | 10 | 4 | 25 | 3 | 1 | 5 | 0.250 | 0.882 |
| 25 | 2.26 | 3.11 | 22 | 0 | 17 | 7 | 0 | 2 | 1.396 | 0.237 |

It is observed that there is a difference between Grade 3A hospitals and non-Grade 3A hospitals only in Sequence generation and related details (item 8), as detailed in Table 3.

In addition, among the 25 items, Ancillary analyses (item 18) had no studies reported, and items 11, 14, and 23 had the highest frequency of being reported as failed.

### Impact of different sizes of hospitals (number of beds), number of authors, sample size, and availability of funding on evaluation quality

The size of the hospital is indicated by the number of established beds in the hospital to which it belongs. Funding support showed whether there was a source of funding for the trial, and authors were asked to report any sources of support for the study, such as project fund sponsored, supply of drugs or equipment, etc. The correlations between the 25 items and the number of beds, number of authors, and sample size are shown in Table 4.

**Table 4.** Correlations between number of beds, authors, sample size, funding and 25 items, total score.

| Item | Number of beds | Number of authors | Sample size | Funding |
| --- | --- | --- | --- | --- |
| 1 | 0.154 | 0.292* | -0.300* | 0.035* |
| 2 | 0.178 | 0.454** | 0.037 | 0.003** |
| 3 | 0.06 | -0.097 | -0.07 | 0.398 |
| 4 | -0.139 | 0.207 | 0.094 | 0.506 |
| 5 | -0.337* | 0.173 | -0.157 | 0.036* |
| 6 | -0.028 | 0.328* | -0.311* | 0.272 |
| 7 | 0.153 | 0.304* | -0.261 | 0.22 |
| 8 | -0.042 | 0.324* | -0.003 | 0.331 |
| 9 | 0.271 | 0.452** | -0.222 | 0.025* |
| 10 | 0.344* | 0.468** | -0.272 | 0.040* |
| 11 | 0.129 | 0.396** | -0.246 | 0.016* |
| 12 | 0.191 | 0.244 | 0.05 | 0.398 |
| 13 | 0.276 | 0.312* | 0.108 | 0.415 |
| 14 | 0.11 | 0.118 | 0.057 | 0.221 |
| 15 | 0.126 | 0.315* | -0.127 | 0.079 |
| 16 | 0 | 0 | 0 | 2 |
| 17 | 0 | 0 | 0 | 2 |
| 18 | 0 | 0 | 0 | 2 |
| 19 | 0.212 | 0.239 | -0.321* | 0.303 |
| 20 | 0.23 | 0.311* | -0.22 | 0.433 |
| 21 | -0.085 | -0.146 | 0.065 | 0.398 |
| 22 | 0.06 | 0.049 | 0.015 | 0.398 |
| 23 | 0.271 | 0.642** | -0.26 | 0.005** |
| 24 | 0.176 | 0.508** | -0.128 | 0.025* |
| 25 | 0.012 | 0.527** | -0.154 | 0.000** |
| Total | 0.254 | 0.598** | -0.254 | 0.000** |
| * p<0.05 |  | ** p<0.01 |  |  |

The number of authors showed a significant correlation with Item1, 2, 6, 7, 8, 9, 10, 11, 13, 15, 20, 23, 24, 25, 14 items and the total score; the sample size had a certain negative correlation with Item1, 6, 19, 3 items; the number of beds established in the affiliated hospitals had a certain negative correlation with Item5, and a certain positive correlation with Item10; the presence of funding showed a certain significant difference for Item1, 2, 5, 9, 10, 11, 23, 24, 25, 9 items and the total score. Scatter plots were drawn with the independent variable as the number of beds and the dependent variable as the total evaluation score, and the total score was not correlated with the size of the hospital to which it belongs as seen in Fig 3 (P>0.05).

**Figure 3.**
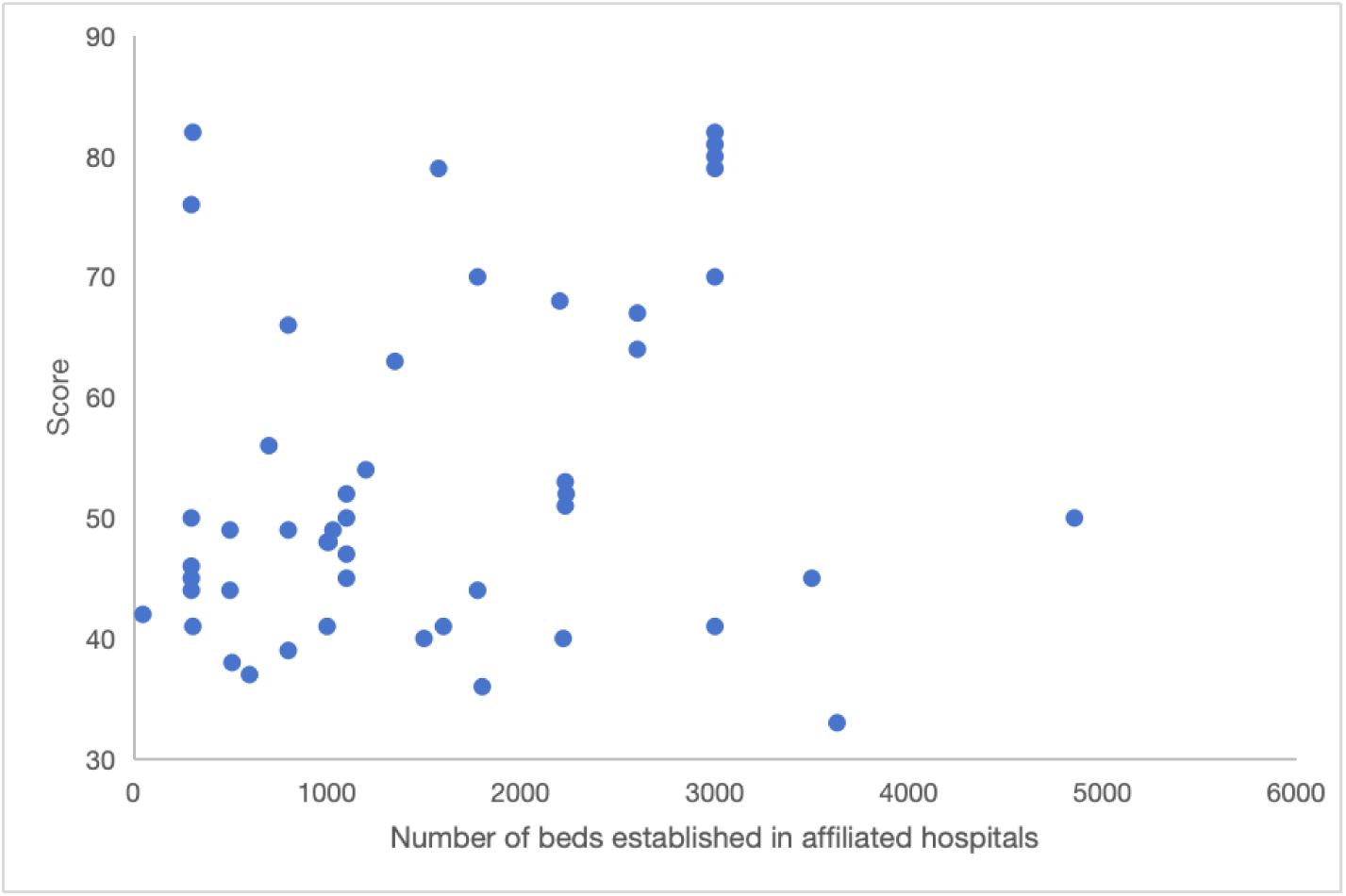
Scatterplot of the number of beds established in affiliated hospitals versus the total score.

### The effect of economic level in different regions on the quality of reports

In Fig 4, The GDP per capita in 2023 of the cities where the hospital the study belongs to is the independent variable, and the total score of the report quality is the dependent variable, inferring from the scatter plot that there is no significant correlation between the total score of the report quality and the level of the economy of the region(P=>0.05).

**Figure 4.**
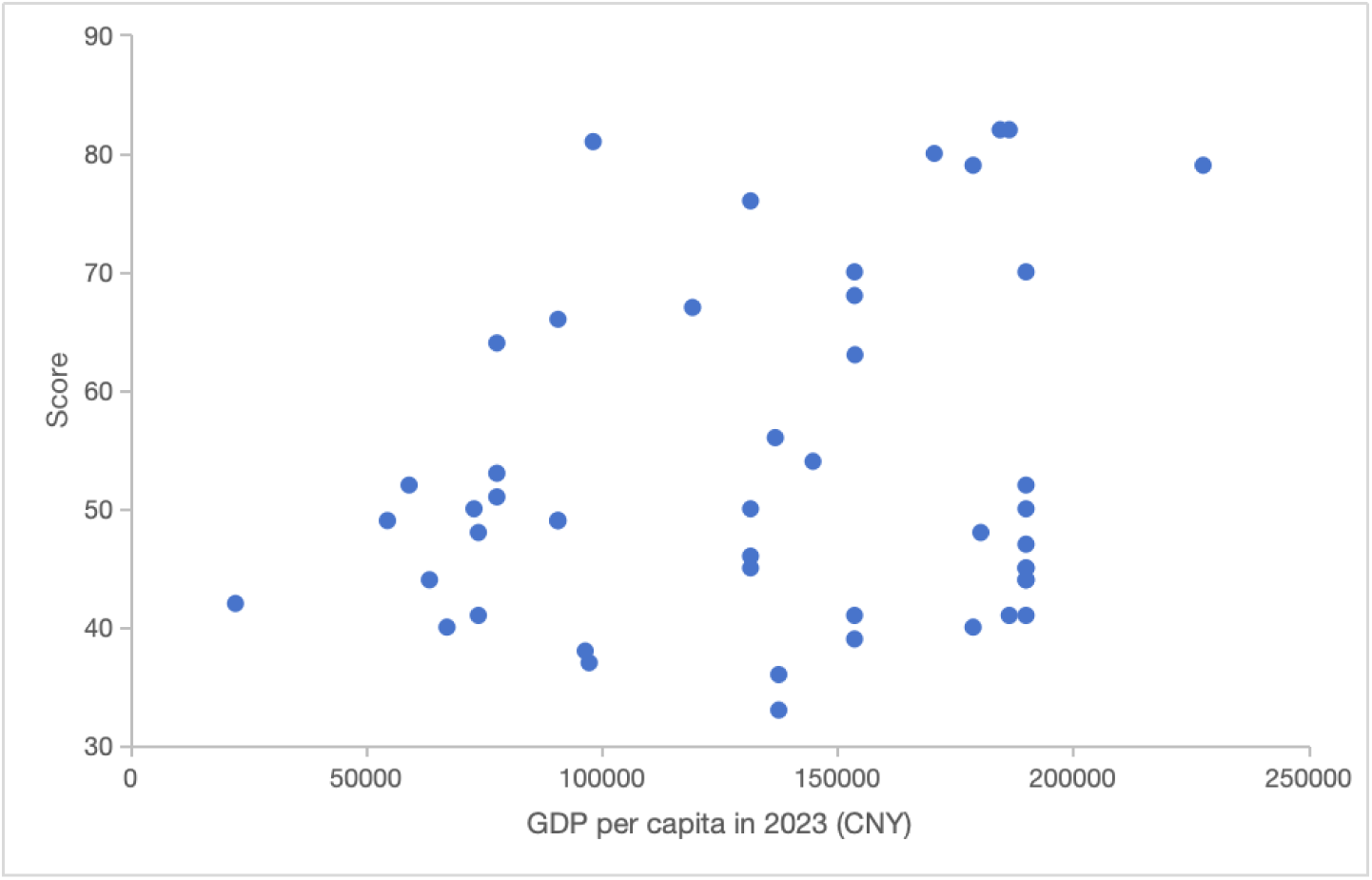
Scatterplot of GDP per capita in 2023 and scores of report quality.

### Multi-factor impact analysis

The Pearson test is shown in Table 5 for a total of six impact factors, including the number of beds established in the hospitals, the grade of the hospitals, the number of authors, the sample size, the presence of funding and the per capita GDP of the affiliated region.

**Table 5.**
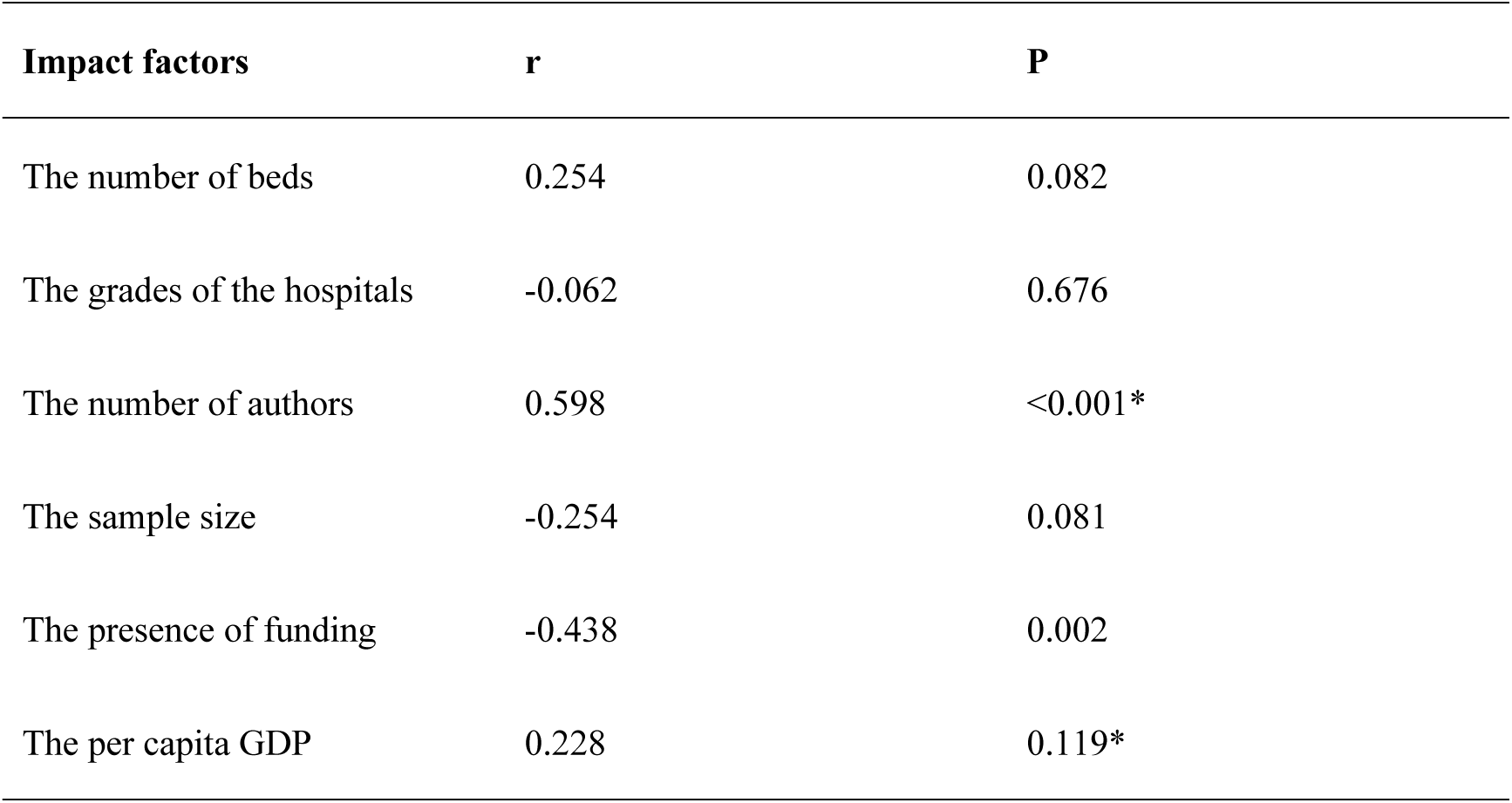
Pearson test for six impact factors.

The number of authors and the existence of foundation funding were the most important factors affecting the report score, and the number of beds prepared by the hospitals, the grade of the hospitals, the sample size, and the per capita GDP of the affiliated regions were not related to the total report score of the evaluation.

## Discussion

This article was the first attempt to evaluate the design quality of acupotomy RCTs according to the CONSORT statement (2010). 48 acupotomy RCTs published in PubMed from January 2006 to January 2024 were evaluated based on the 25 items that were evenly scored. The previous studies focusing on acupotomy mostly were systematic reviews and Meta-analyses, with risk of bias as the principle for evaluating overall quality(41-45), lack of studies using the CONSORT checklist to assess the quality of acupotomy RCTs, which was met by this study.

This article used the international RCT standard CONSORT guidelines, which are more authoritative for quality evaluation(46, 47). In addition, in studies using CONSORT for evaluating the quality of RCTs, most only selected part of 25 items(48-50). But in this article, the 25 items of CONSORT were averaged scored, with the same weighting between the items, which emphasized more on the completeness and standardization of the article’s structure and content. Therefore, the standardization and comprehensiveness of data is better.

### Overall quality to be improved

In studies of acupotomy RCTs from 2006 to 2024, the overall quality needs to be improved. Among the 48 studies, the lowest score was 33 points, the highest score was 82 points, the average score was 53.1 points, and the median score was 49 points. This condition is not uncommon, and a 2021 Korean study similarly showed that the reports’ quality of acupotomy therapy is currently low(51).

As shown by some studies, there is still no significant improvement in the quality of RCT reports in fields such as dentistry, psychiatry, psychology, cardiothoracic surgery, etc., since the release of the updated CONSORT list in 2010(52-56), Qi Cui MM et al. also argued that the quality of current RCTs is not as great as expected(57), and the mere publication of the CONSORT guidelines did not appear to have a significant impact on the quality of RCTs, whereas the authors’ awareness of the CONSORT items was relevant to the improvement of the RCTs quality(52, 58, 59).

Since factors such as the training background of the researchers, the original design of the trial, and the requirements of the journals are closely related to the quality of the final report, improving the design, implementation, and reporting of RCTs is a systematic and multifaceted task that demands a very close understanding of and rigorous compliance with the CONSORT checklist to ensure RCTs’ completeness and standardization, systematically accomplishing the entire experimental procedure(57, 60).

### Different quality levels among different items

There were large differences in quality levels between 25 items, and some items were still irregularly reported. The Ancillary analyses were not mentioned in any of the articles, and Blinding, Recruitment, and Registration were the three items with the lowest quality of reporting, which probably was due to the lack of rigor in blinding methods, poor recruitment methods, and omission of key information(61, 62).

It is one of the key features of RCTs that lack of blinding may lead to a risk of bias in treatment effects(63). Although there was previously some controversy over the method of blinding(64), the researchers suggest that the integrity of blinding must still be maintained by devoting sufficient resources, consideration, and programming to it(65).

Recruitment of physicians and patients has always been one of the difficulties in clinical trials(66), most trials fail to complete recruitment on time, resulting in longer recruitment time, less reliable results, and even additional costs (67). Therefore, some scholars evaluated the potential factors affecting the effectiveness of recruitment, which revealed that better recruitment methods, as well as monitoring and managing poor recruitment, can contribute to the success of recruitment(68).

Similar to the International Conference on Harmonisation of Technical Requirements for Pharmaceuticals for Human Use(ICH), whether or not a study is registered is an important factor in study reliability(69).

A previous study also found fewer reports of methodological quality in the areas of randomization methods, allocation concealment, and blinding details(70-73), probably because methodological content has been analyzed separately from other CONSORT projects previously(74, 75).

Therefore, based on the updated CONSORT guidelines, researchers should focus on the overall structural integrity of the design quality of RCTs.

### Higher potential for use of CONSORT in Grade 3A hospitals

This study showed that over 80% of the 48 studies were from Grade 3A hospitals, which shows that in studying the efficacy of Acupotomy, a larger percentage of hospitals with higher grades are among those that follow the CONSORT statement. Such hospitals have more stringent requirements for the process of randomized controlled trials, which may be due to the more advanced equipment in higher-grade hospitals, and a better-quality clinical trial environment provides hospitals with more accurate data sources and results, which is consistent with the findings of Xuan Zhang and other scholars(76).

Nisha Berthon-Jones et al. proposed that multicenter randomized clinical trials facilitate meeting required timelines and increasing cost-effectiveness by initiating, recruiting, and collecting data and samples in different areas(77), so that, high-quality multicenter RCTs can strengthen the external validity of findings in practice(78).

But among 25 items, the parts of Sample size (item7), Allocation concealment mechanism (item9), Implementation (item10), Blinding (item11), Recruitment (item14), Ancillary analyses (item18), Harms (item19) are poorly reported by these articles, which means that the hospitals should pay more attention to the details.

We found that in the Ancillary analyses (item18) section, the compliance of both Grade 3A hospitals and non-Grade 3A hospitals was not enough. The investigators should state the results of the study completely in the report so as not to affect the results of the trial or lead to duplication and waste of future research, and how to improve the quality of the report further was all mentioned in these reports. This view was echoed by Mercieca-Bebber R et al.(79).

In addition, as hierarchical medical system continues to advance in China, the relevant authorities can introduce policies to encourage cooperation and exchanges between hospitals in different grades, clarify the roles and priorities of their respective clinical fields, and realize the rational allocation of high-quality medical resources, thereby taking advantage of each hospital’s strengths, promoting the conduct of high-quality clinical trials, improving the quality of research, and strengthening the scientific research capacity of the hospitals(80, 81).

### Trend of increasing scores over time

In the 18 years since the first small-needle RCT was published in 2006(82), the quality of articles has gradually improved over time. This is consistent with the study by Fay Karpouzis et al. suggesting that the report quality of the medical literature has significantly improved with the adoption of the CONSORT guidelines(83-85). The CONSORT statement was first published in 1996(86), and officially translated and endorsed by the CONSORT panel in 2007. Scholars such as Tian-Jiao Song have shown that although CONSORT has been neglected during this period, the evidence suggests that the overall quality of RCTs published in Chinese journals has improved from 2005 to 2015(31, 60, 87).

On the one hand, with the development of science and technology, the medical equipment and technology in clinical experiments have been improving, and a favorable medical experimental environment is a necessary prerequisite for research results; on the other hand, with the constant refinement of the preliminary experiments, it also provides abundant theoretical and practical experience for the later experiments, therefore the quality of the experimental reports has been improving with the time.

According to Letícia Maria Wambie, the increase in scores on the CONSORT assessment reflects the growing familiarity of researchers with the rules of CONSORT, which in turn improves their research reporting skills(88), and the CONSORT checklist is constantly updated, with statements becoming clearer and more accurate, which will also contribute to the quality of reporting(12, 89), to some extent reflecting the mutually reinforcing relationship between the two.

### Factors affecting the design quality of RCTs

Among the factors affecting the report quality, the number of authors and the presence of fund support had a certain impact on most of the evaluation indicators, while the sample size and the number of established beds in the affiliated hospitals had an impact on only a few of the indicators, and whether it was a Grade 3A hospitals, the economic level of the region and the quality of the report had no correlation with each other. Sabapathy P. Balasubramanian et al. similarly found high number of authors, multicenter studies, and statement of funding sources to be factors significantly associated with better quality of reporting in a general surgery RCT quality assessment(90, 91).

This suggests that the size of the research team and financial support are important for improving the quality of reporting, but simply increasing sample size and hospital size does not completely address the issue of reporting quality(92). Furthermore, even if it is a large hospital or a more economically developed region, it does not mean that all of its studies are of high quality. This finding is partially consistent with the findings of Ioannis Liampas et al. who concluded that the number of authors had a meaningful impact on the study, whereas sample size and commercial funding reports had a less impact on the study(93). A study by Parish AJ et al. concluded that scientific collaboration (number of authors) is associated with higher citation impact(94). Although the primary responsibility for improvement rests with the investigators(95), reviewers and editors of surgical journals can promote this process by endorsing CONSORT guidelines(96, 97), such as requiring authors to follow the guidelines as a condition of publication, and assessing the reliability of RCTs to be published(98).

Therefore, advocating more efficient cooperation and resource sharing, such as cross-region, cross-hospital, and multi-team cooperation, is conducive to integrating the resources of all parties, and sharing the experience of trial design, data collection, and statistical analysis, to improve the feasibility and efficiency of the trial, which will establish a more regularized standard of research based on consensus(99, 100).

In further study, the number of included articles, the weight allocation of items in CONSORT and the analysis of CONSORT should be improved. The number of included articles needs to be raised, for one is due to the international terminology of acupotomy is not clear, and the words used in previous studies are confusing, this study has tried to check the whole, but there may still be omitted articles, the second is that this study lacks the articles of 2010, 2015, which may affect the analysis. The weights of 25 items need to be clarified, there were sub-items for different items, with different content and length, which were of different significance for the article, affecting the different value evaluation of the RCTs from different perspectives. The instructions of CONSORT need to be refined, the CONSORT instructions only have short statements currently, which are understood differently by people with different research backgrounds and experiences, it is recommended that examples or detailed instructions be introduced(101). Due to the limited sample, there are limited statistical analyses available, and it is hoped that with the inclusion of more relevant literature and more standardized evaluation, a more in-depth analysis of influencing factors can be carried out.

## Conclusion

This study showed that of the 48 Acupotomy RCT studies published during 2006-2024, based on the CONSORT statement, the highest score was 82, the lowest score was 33, the mean score was 53.1, and the median score was 49, and there was a positive correlation between the report quality and the time of publication.

Among the many factors affecting the quality of the report, the number of authors and the possession of foundation support were the most important factors affecting the total score, while the number of beds in the affiliated hospital, the hospital grade, the number of samples, and the per capita GDP of the affiliated region did not correlate with the total score of the report assessment. Among the 25 items, Ancillary analyses were not reported by any study, as well as Blinding and Recruitment were reported as failed with the highest frequency.

Therefore, updating and standardizing the use of the CONSORT checklist can help to improve the quality of RCT reports, and cross-team communication, cooperation and resource sharing can help to integrate resources and establish a more standardized research standard.

## Data Availability

All relevant data are within the manuscript and its Supporting Information files.

## Acknowledgements

We would like to thank three research funds for their support.

## List of abbreviations

RCTs: Randomised controlled trials
CONSORT: Consolidated Standards of Reporting Trials
TCM: Traditional Chinese Medicine
GDP: Gross Domestic Product
CAM: Complementary and Alternative Medicine
NK-F: Needle-knife fistulotomy

## Notes

### Competing Interest Statement

The authors have declared no competing interest.

### Clinical Trial

N/A This article is an general evaluation of 48 published clinical reports that have been published within ethic approvals. The dates in this article were collected from these reports, which were published and ethically approved. Therefore, this article not involving clinical trails personally, and the ethics approval of human participants research.

### Funding Statement

Yes

### Author Declarations

N/A This article is a general evaluation of 48 published clinical reports that have been published within ethic approvals. The dates in this article were collected from these reports, which were published and ethically approved. Therefore, this article not involving clinical trails personally, and the ethics approval of human participants research.

